# Muscle and Adipose Tissue Segmentations at the C3 Vertebral Level for Sarcopenia-Related Clinical Decision-Making in Patients with Head and Neck Cancer

**DOI:** 10.1101/2022.01.23.22269674

**Authors:** Kareem A. Wahid, Brennan Olson, Rishab Jain, Aaron J. Grossberg, Dina El-Habashy, Cem Dede, Vivian Salama, Moamen Abobakr, Abdallah S.R. Mohamed, Renjie He, Joel Jaskari, Jaakko Sahlsten, Kimmo Kaski, Clifton D. Fuller, Mohamed A. Naser

**Author notes:** co-corresponding authors Corresponding author’s contact information: Clifton D. Fuller. Mohamed A. Naser. Postal Address: The University of Texas MD Anderson Cancer Center, 1515 Holcombe Blvd, Houston, TX, 77030, USA.

## Abstract

The accurate determination of sarcopenia is critical for disease management in patients with head and neck cancer (HNC). Quantitative determination of sarcopenia is currently dependent on manually-generated segmentations of skeletal muscle derived from computed tomography (CT) cross-sectional imaging. This has prompted the increasing utilization of machine learning models for automated sarcopenia determination. However, extant datasets of head and neck CT imaging currently do not provide the necessary manually-generated skeletal muscle segmentations at the C3 vertebral level needed for building these models. In this data descriptor, we detail the annotation of a large set of head and neck CT images for use in automated sarcopenia-related clinical decision making and body composition analysis. A set of 394 HNC patients were selected from The Cancer Imaging Archive, and their skeletal muscle and adipose tissue was manually segmented at the C3 vertebral level using sliceOmatic in .tag format. Subsequently, using publicly disseminated Python scripts, we generated corresponding segmentations files in Neuroimaging Informatics Technology Initiative format. In addition to segmentation data, additional clinical demographic data germane to body composition analysis have been retrospectively collected for these patients from the University of Texas MD Anderson Cancer Center databases. These data are a valuable resource for studying sarcopenia and body composition analysis in patients with HNC.

## Background & Summary

Head and neck cancer (HNC) affects more than 900,000 individuals worldwide annually ^1^. Sarcopenia, a body composition status describing skeletal muscle depletion, is a well-validated negative prognostic factor in patients with HNC and has become increasingly studied in recent years ^2–4^. Sarcopenia is quantitatively determined primarily using the cross-sectional estimate of skeletal muscle at a specific vertebral level. Current methods to generate cross-sectional skeletal muscle segmentations for use in sarcopenia determination are reliant on expert human-generated segmentations, which can be time-consuming to procure and subject to user variability ^5^. Therefore, the dissemination of high-quality skeletal muscle segmentations is of paramount importance to develop tools for sarcopenia-related clinical decision making.

Publicly disseminated HNC datasets have increased sharply in recent years. For example, several HNC imaging datasets, predominantly composed of computed tomography (CT) images, have been hosted on The Cancer Imaging Archive (TCIA) ^6^. Public datasets, such as these, have been crucial towards advanced algorithmic development for clinical decision support tools ^7^. However, only a handful of existing HNC datasets have provided information germane to determining sarcopenia status in patients, namely that by Grossberg at el. ^8^ providing body composition analysis data based on abdominal imaging. Moreover, to date, there are no existing open-source repositories for body composition analysis data based on head and neck region imaging. Increasing evidence has shown the potential utility of sarcopenia determination using skeletal muscle in the head and neck region ^2,9^. This is driven by the fact that many patients with HNC may not have abdominal imaging as part of the standard workup, but will almost certainly have head and neck region imaging, particularly due to its requirement for radiotherapy treatment planning ^10^ and staging purposes ^11^. These head and neck imaging data could be used to train models for automated sarcopenia-related clinical decision making, as shown in previous studies ^12^. Therefore, the dissemination of sarcopenia-related data derived from head and neck imaging is an unmet need for the research community that may foster more rapid adoption of automated HNC clinical decision support tools.

Here we present the curation and annotation of a large-scale TCIA dataset of 394 patients with HNC for use in sarcopenia-related clinical decision making and body composition analysis. The primary contribution of this dataset is high-quality skeletal muscle and adipose tissue segmentation at the cervical vertebral level in an easily accessible and standardized imaging format, in addition to additional clinical demographic variables. These data can be leveraged to build models for body composition analysis and sarcopenia-related decision-making germane to HNC. Moreover, these data could form the basis for future data modeling challenges for sarcopenia-related decision-making in patients with HNC. An overview of the data descriptor is shown in **Figure 1**.

**Figure 1:**
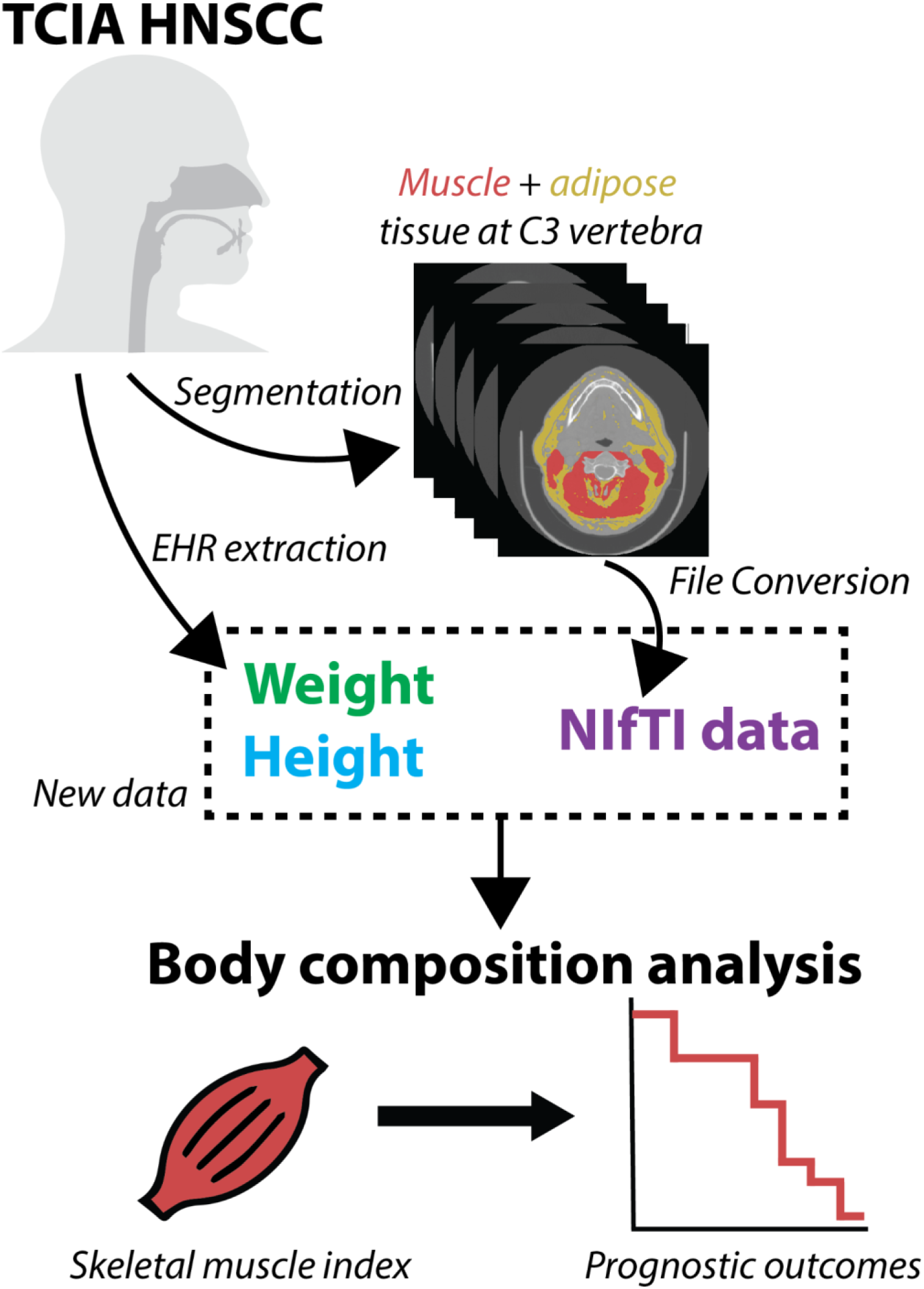
Data descriptor overview. The Cancer Imaging Archive (TCIA) head and neck squamous cell carcinoma (HNSCC) computed tomography dataset is used to generate muscle and adipose tissue segmentations at the third cervical (C3) vertebral level in Neuroimaging Informatics Technology Initiative (NIfTI) format. Additional demographic data (weight, height) is collected from electronic health records (EHR). The final newly distributed dataset can be used for body composition analysis, such as sarcopenia-related clinical decision-making.

## Methods

### Study Population and Image Details

To develop this dataset, imaging data from the TCIA head and neck squamous cell carcinoma (HNSCC) collection, a large repository of imaging data originally collected from The University of Texas MD Anderson Cancer Center, were utilized. Specifically, 396 patients with contrast-enhanced CT scans were selected from the 495 available patients in the “Radiomics outcome prediction in Oropharyngeal cancer” dataset ^13,14^. These patients were selected due to their inclusion of the third cervical vertebral level on imaging. To summarize the underlying data, these were patients with histopathologically-proven diagnosis of squamous cell carcinoma of the oropharynx that were treated with curative-intent intensity-modulated radiotherapy. Imaging data was composed of high-quality CT scans of patients who were injected with intravenous contrast material. Images were acquired before the start of radiotherapy. Imaging data were provided in the Digital Imaging and Communications in Medicine (DICOM) standardized format. Additional details on the original imaging dataset are provided in the corresponding data descriptor ^14^ and TCIA website ^13^. All DICOM images were previously de-identified, as described in previous data descriptors ^8,14^.

### Skeletal Muscle Segmentation

For each CT image, the middle of the third cervical vertebra (C3) was located on a single axial slice and the muscle and fat tissues were manually segmented. As described in previous publications ^15^, muscle and fat tissue were defined in the ranges of -29 to 150 and -190 to -30 Hounsfield units, respectively. Paraspinal and sternocleidomastoid muscles were segmented as previously described, while adipose tissue segmentation included subcutaneous, intermuscular, and visceral compartments. Manual segmentations were performed using a commercial image-processing platform (sliceOmatic v. 5.0, Tomovision). Examples of skeletal muscle and adipose tissue segmentations with corresponding images are shown in **Figure 2**. Segmentations were exported from sliceOmatic in .tag format, with the corresponding 2D axial slice in DICOM format.

**Figure 2:**
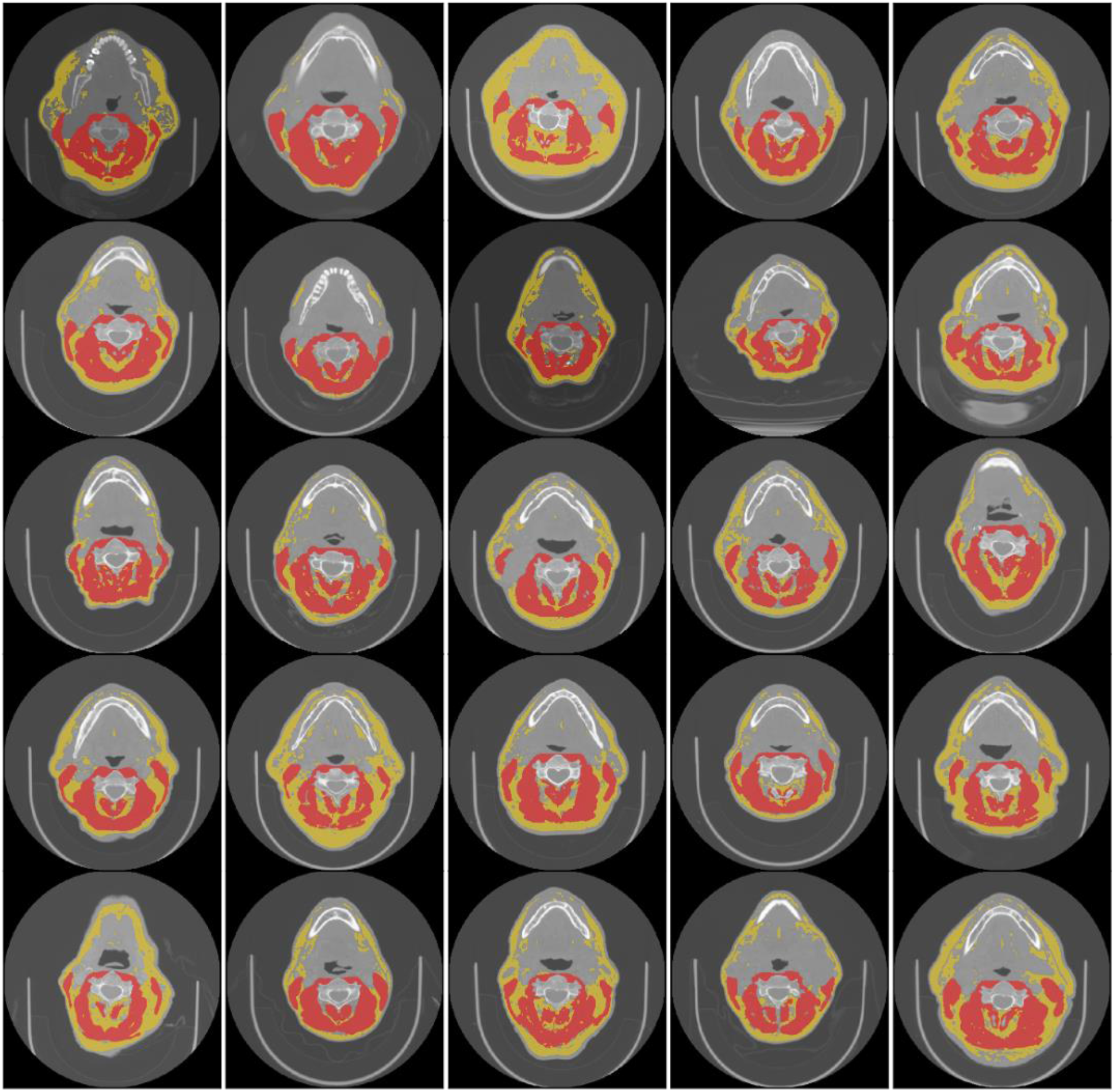
Segmentation examples for a subset of 25 cases. Each image corresponds to one patient. Images are single-slice computed tomography axial views with segmentations superimposed. The red regions correspond to skeletal muscle tissue and the yellow regions correspond to adipose tissue.

### NIfTI Conversion

The Neuroimaging Informatics Technology Initiative (NIfTI) file format is increasingly seen as the standard for reproducible imaging research ^16^. Therefore, we converted all our segmentation (.tag) and imaging (.dcm) data to NIfTI format, in order to increase the interoperability and widespread utilization of these data.

For all file conversion processes, Python v. 3.7.9 ^17^ was used. An overview of the NIfTI conversion workflow for segmentations and images is shown in **Figure 3**. In brief, using an in-house Python script, .tag files were read in binary format and converted into numpy format ^18^, trimmed to remove header information, and then re-sized to the corresponding size of the 2D DICOM axial slice (also in numpy format), i.e., a 2D array. The slice location was determined from the 2D DICOM axial slice in tandem with the 3D DICOM image (acquired from the TCIA) using pydicom ^19^. A 3D array, that contained the segmentation information, was then created by filling in all non-segmented slices with 0s, yielding a 3D segmentation mask. Each non-zero entry, corresponding to separate regions of interest in the 3D segmentation mask (1 = muscle, 2 = adipose for toy example in **Figure 3**), was then converted to binary masks in NIfTI format (individual files for each region of interest) using simpleITK ^20^. 3D CT DICOM images were loaded into python using the DICOMRTTool ^21^ library, and then converted to NIfTI format using simpleITK. Additional documentation on scripts used for conversion can be located on the corresponding GitHub repository: https://github.com/kwahid/C3_sarcopenia_data_descriptor.

**Figure 3:**
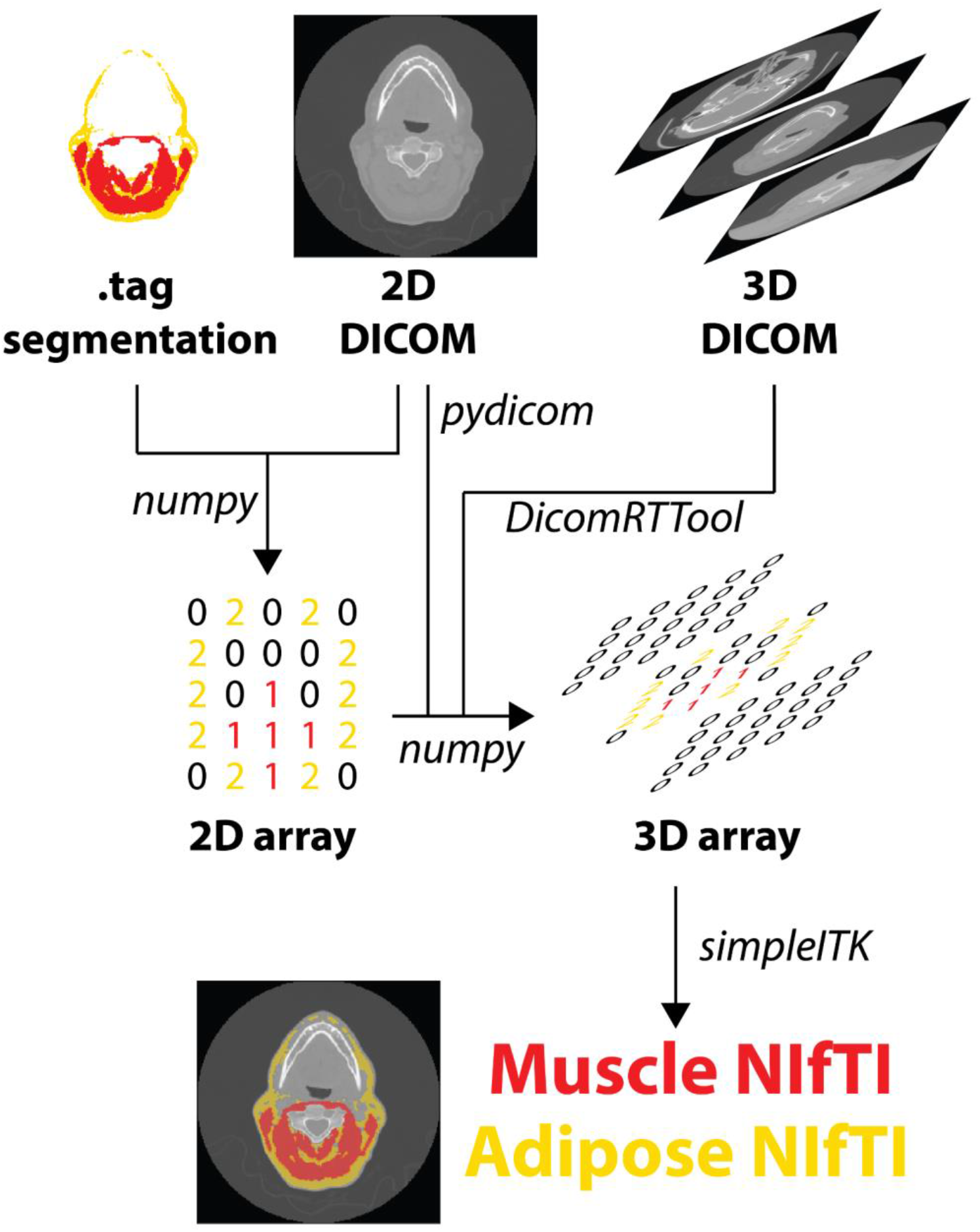
File conversion workflow for segmentations and images. Outputs from sliceOmatic software, i.e., .tag segmentation and 2D Digital Imaging and Communications in Medicine (DICOM) slice, are used to generate a 2D mask array of muscle and adipose tissue. Information from 2D DICOM slice and corresponding 3D DICOM image (acquired from corresponding The Cancer Imaging Archive dataset) are used to generate a 3D array, which is then converted to Neuroimaging Informatics Technology Initiative (NIfTI) format.

Of the 396 cases converted through the previously mentioned workflow, one patient (TCIA ID 0435) had a DICOM CT file with image reconstruction errors, while another (TCIA ID 0464) was unable to be converted to NIfTI format successfully, that necessitated their removal from the final dataset, yielding 394 image/segmentation pairs in NIfTI format.

### Additional Patient Demographic Data Collection

In addition to cross-sectional area derived from skeletal muscle segmentations, calculation of skeletal muscle index requires data concerning patient height. In order to increase the usability of segmented regions of interest for use in sarcopenia-related calculations and model building, we also collected corresponding height (in m) and weight (in kg) data for all patients in our dataset. Anonymized TCIA IDs were mapped to existing patient medical record numbers to collect the corresponding data. Data were collected from the University of Texas MD Anderson Cancer Center clinical databases through the EPIC electronic medical record system by a manual review of clinical notes and paperwork. The Institutional Review Board of the University of Texas MD Anderson Cancer Center gave ethical approval for this work (RCR03-0800). Height and weight were collected for the pre-radiotherapy visit only in accordance with the pre-radiotherapy imaging collected for this study. Clinical data collection was performed by a trained physician (D.E.).

### Data Records

#### Segmentation Data

This data collection consists of 788 3D volumetric compressed NIfTI files (394 skeletal muscle “muscle.nii.gz” files, 394 adipose tissue “fat.nii.gz”) derived from an original collection of 394 DICOM files of pre-therapy CT images collected from 495 patients originally on the TCIA (“Radiomics outcome prediction in Oropharyngeal cancer”) ^13,14^. The skeletal muscle and adipose tissue NIfTI files are binary masks (0 = background, 1 = region of interest). While we do not provide the corresponding 394 CT images in nifti format due to Figshare upload size constraints, we do provide all the code necessary to produce these files (see code availability section). In addition to NIfTI format files, we also include .tag segmentation files and corresponding 2D DICOM files (sliceOmatic outputs) for interested parties to recreate our NIfTI conversion pipeline if desired. Of note, we do not include the 3D DICOM CT files as these can be acquired from existing TCIA repositories ^13,14^.

#### Clinical Data

We also provide a single comma-separated value (CSV) file containing additional clinical demographic data germane to sarcopenia clinical-decision making. In the CSV file, in addition to newly collected height and weight variables, we also include previously publicly available clinical variables in the TCIA dataset ^13,14^ relevant for body composition analysis (age and sex).

Segmentations are organized by an anonymized TCIA patient ID number (“TCIA Radiomics ID”) and can be cross-referenced against the CSV data table using this identifier. The data records and supplemental descriptions of the meta-data files are cited under Figshare: 10.6084/m9.figshare.18480917.

### Technical Validation

#### Skeletal muscle segmentations

The segmentations provided in this data descriptor have been utilized as ground-truth segmentations in a previous study by Naser et al. ^12^ which yielded sarcopenia determination results (normal vs. depleted skeletal muscle) that were consistent with existing literature ^9^, i.e., overall survival stratification is significant in males but not females as determined by Kaplan Meier analysis. Note: 4 patients included in the current data descriptor were excluded from the aforementioned analysis (TCIA ID’s: 0226, 0280, 0577, and 0607), due to oblique imaging orientations.

#### EPIC (Electronic Medical Record System)

The University of Texas MD Anderson Cancer Center adopted this system in the year 2017 which allows integrating research data and accessing data from virtually every electronic source within the institution. https://www.clinfowiki.org/wiki/index.php/Epic_Systems.

### Usage Notes

This data collection is provided in NIfTI format with the accompanying CSV file containing additional clinical information indexed by TCIA identifier. We invite all interested researchers to download this dataset to use in sarcopenia-related research and automated clinical decision support tool development.

Images (reproducible through code) and segmentation are stored in NIfTI format and may be viewed and analyzed in any NIfTI viewing application, depending on the end-users requirements. Current open-source software for these purposes includes ImageJ ^22^ and 3D Slicer ^23^.

## Data Availability

All data produced are available online at Figshare (doi: 10.6084/m9.figshare.18480917).

## Code Availability

Segmentation was performed using the commercially-available tool sliceOmatic v. 5.0 (Tomovision).

The code for NIfTI file conversion of DICOM CT images and corresponding .tag format muscle/adipose tissue segmentations was developed using in-house Python scripts and is made publicly available through GitHub: https://github.com/kwahid/C3_sarcopenia_data_descriptor. Alternative code for converting .tag files to Matlab readable format can be located at: https://github.com/RJain12/matlab-tag-reader.

